# The Pediatric Sleep Questionnaire as a screening tool for sleep-related breathing disorders: An Umbrella Review

**DOI:** 10.1101/2024.05.14.24307375

**Authors:** David Okuji, Amanda Newcity, Amir Yavari, Farnaz Shahraki, Oghenerukevwe Erhenede, Qaiser Ahmed, Rachel Patenaude, Alberta Twi-Yeboah, Richard McGowan

## Abstract

**Purpose:** The purpose of this umbrella review was to assess available systematic reviews and meta-analyses reporting on the use of the Pediatric Sleep Questionnaire as a screening tool for identifying sleep related breathing disorders in children, with subsequent referral from dentists to physicians.

**Methods:** A comprehensive literature search was conducted with electronic databases to identify systematic reviews and meta-analyses reporting on the Pediatric Sleep Questionnaire as a screening tool for pediatric sleep related breathing disorders. Eligible reviews were systematically selected and were assessed qualitatively with the AMSTAR 2 tool and quantitatively with meta-analyses methods.

**Results:** A total of 129 articles for possible inclusion were identified, and five studies were selected for quantitative analysis. Of these five studies, the overall AMSTAR 2 quality assessment showed two with moderate and three with low quality ratings. The five studies showed point-estimate mean values for prevalence, sensitivity, specificity, positive predictive value, positive predictive value, negative predictive value as, respectively, respectively, 50.58, 74.97, 49.68, 58.16, 62.24 percent, and the diagnostic odds ratio as 6.94 percent for the Pediatric Sleep Questionnaire tool.

**Conclusions:** Dentists should adhere to the American Academy of Pediatric Dentistry guidelines and know that there is low to moderate evidence to support the utilization of the Pediatric Sleep Questionnaire as a sufficiently accurate screening tool for the diagnosis of sleep related breathing disorders in children, followed by an appropriate referral to a physician for comprehensive assessment and management.

## INTRODUCTION AND RESEARCH QUESTION

### Rationale

Pediatric sleep disorders include sleep issues that meet diagnostic criteria for obstructive sleep apnea, parasomnias, narcolepsy, and insomnia.^1^ These disorders often go undiagnosed and are persistent in children, leading to increased hospital visits, brain damage, seizures, coma, and cardiac complications.^2–5^ It has been reported that up to 40 percent of children experience a sleep problem sometimes between infancy and adolescence.^6^ In 2006, the Institute of Medicine (now known as the National Academy of Medicine) published its profound and probing report, "Sleep Disorders and Sleep Deprivation: An Unmet Public Health Problem".^2^ The key conclusions of this seminal report illuminate an opportunity for dentists to join the medical community for integrative care of children with sleep-related breathing disorders (**SRBDs**). The report specifically notes that there is high demand for the care of children with sleep-related breathing disorders, a great shortage of health care providers to diagnose and treat these children, and recommends an interdisciplinary approach, which involves dentistry in conjunction with other medical and health care domains.

The American Academy of Pediatrics recommends that “all children/adolescents should be screened for snoring” and “if polysomnography is not available, then alternative diagnostic tests or referral to a specialist for more extensive evaluation may be considered.^4^”

The American Academy of Pediatric Dentistry (**AAPD**) guidelines, to which all dentists who treat children should adhere; focus on the screening, clinical assessment, non-surgical intraoral appliances, and referral to medical specialists for patients. Specifically, the guidelines indicate that pediatric patients should be screened for “sleep-related breathing disorders such as obstructive sleep apnea and primary snoring.^5^ “The guidelines suggest nine sleep questions for inclusion on health history forms to assist with the identification of children at risk. Notably, eight of the nine suggested questions are included in the Pediatric Sleep Questionnaire (**PSQ**) screening tool.^7^ As well, the guidelines state that the PSQ is “not sensitive enough to detect presence or severity of OSA.^5^ Thus, it is of critical importance that dentists who treat children conduct a thorough history, clinical evaluation, and screening for sleep-related breathing disorders since they are uniquely positioned to identify children with the greatest risk at semi-annual periodic examination visits.^8^ Two studies showed wide ranges of results for the use of screening tools by pediatric dentists, which estimated 40.7 to 70.5 percent of pediatric dentists routinely screened patients for obstructive sleep apnea, and 4.7 to 39.2 percent utilizing survey questionnaires as the screening tool.^9,10^ This evidence indicates that a minority of pediatric dentists routinely utilize survey questionnaires as screening tools for sleep-related breathing disorders.

Finally, it is critical to distinguish between the use of screening versus diagnostic accuracy metrics. Guidance for high value diagnostic testing suggests that the sensitivity plus specificity should be “at least 1.5.^11^” For screening metrics, “sensitivity and specificity should usually be applied only in the context of describing a screening test’s attributes relative to a reference standard and predictive values are more appropriate and informative in actual screening contexts. Sensitivity and specificity can be used for screening decisions about individual people if they are extremely high. Predictive values need not always be high and might be used to advantage by adjusting the sensitivity and specificity of screening tests. In screening contexts, researchers should provide information about four metrics (sensitivity, specificity, positive predictive value, negative predictive value) and how they were derived and clinical providers should have the skills to interpret those metrics effectively for maximum benefit to patients and the healthcare system.^12^”

### Objectives

The purpose of this study was to conduct an umbrella systematic review and determine the utility of the pediatric sleep questionnaire as a screening tool for SRBDs in children, based upon the application of diagnostic test accuracy metrics for use as a screening, rather than, a diagnostic tool. The rationale is to provide guidance to dentists who treat children to use the PSQ as a screening tool to screen children for SRBDs. The main research question for this umbrella review is: “For children under age eighteen years old is the PSQ sufficiently accurate as a screening tool for to estimate a diagnosis of SRBDs, for referral to physicians by dentists, when compared to the gold standard polysomnography test (**PSG**)?” The hypothesis is the PSQ is sufficiently accurate as a screening tool for the diagnosis of SRBDs in children.

## METHODS

### Research Protocol and Registration

The umbrella review protocol was registered on January 19^th^, 2023, and assigned the identification number CRD42023393232 in the PROSPERO^13^ international prospective register of systematic reviews hosted by the National Institute for Health Research, Center for Reviews and Dissemination, University of York, UK.

### Reporting format

The Preferred Reporting Items for Systematic Reviews and Meta-analysis (PRISMA) was used throughout the process of this umbrella review.^14^

### Eligibility criteria

#### Population, Intervention, Comparator, Outcome (PICO)

The population included children under age 18 years old, regardless of sex, race, socioeconomic status, health status, or geographic location. The indicator was all child-subjects were screened with the PSQ tool. The comparator was all children underwent PSG testing. The outcomes were an Apnea Hypopnea Index (**AHI**) score greater than or equal to one.

##### Inclusion Criteria

The reviewed articles were designed as either a systemic review or meta-analysis, which included the PSQ as a screening tool and the PSG as a diagnostic tool. The subjects included in the study were children less than 18 years old, regardless of health status.

##### Exclusion Criteria

Articles which were not designed as systematic review with meta-analysis were excluded from the study even if they utilized PSQ as a screening tool or PSG as a diagnostic tool. Articles which were classified as case reports, comments, laboratory studies, letters, and narrative reviews were excluded.

### Literature Search

#### Search Strategy, Information Sources, and Selection Process

An initial literature search was conducted on March 18, 2022, in all relevant publications in the PubMed, Embase, Cumulative Index to Nursing & Allied Health (**CINAHL**), Scopus, Web of Science, Cochrane Database of Systematic Reviews, Dentistry & Oral Sciences Source (**DOSS**) and, Health and Psychosocial Instruments (**HaPI**) databases, as well searching the grey literature. Hand searching and checking reference lists were also used to identify relevant additional records. The search strategy was composed of the following keywords and Boolean operators "pediatric sleep questionnaire" or “PSQ” and applied filters for “meta-analysis,” “review,” “systematic review.” An updated search was conducted on June 13, 2023, to identify additional studies.

### Data Collection and Analysis

#### Study selection

For the study selection, two authors independently screened the titles and abstracts with the Covidence platform to remove duplicates and read the full text of all papers to identify relevant systematic reviews, with meta-analyses. Discrepancies with selected reviews were resolved through discussion and mutual agreement by the two researchers.

#### Synthesis Methods and Data Extraction

##### Synthesis Methods

The methods utilized to synthesize the data from the reviewed articles included the compilation of characteristics of the included studies, corrected covered area (**CCA**) degree of overlap, effect size of variables from the meta-analysis, “A MeaSurement Tool to Assess systematic Reviews” (**AMSTAR**) 2 quality appraisal, risk of bias, heterogeneity of the data, and certainty of evidence (**COE**).

##### Data collection process and Data items

Qualitative data extraction for the characteristics of the included systematic review and meta-analysis studies was completed by two of this study’s authors who independently focused on collection details on the diagnostic tests, author names, publication year, journal name, research question, search strategies, study design, outcomes, assessment tools, and conclusions for each study marked for inclusion in this investigation. Disagreements were resolved through discussion and mutual agreement by the two researchers.

Quantitative data extraction was completed by the same two independent reviewers. For the meta-analysis summary for this part of the present study, the author names, year of publication, number and study design of the primary studies, specific diagnostic test estimates with confidence intervals, heterogeneity statistics and quality appraisal, with the AMSTAR 2 tool, were collected.

The quantitative data included prevalence, sensitivity (**SE**), specificity (**SP**), positive predictive value (**PPV**), negative predictive value (**NPV**), positive likelihood ratio (**LR+**), negative likelihood ratio (**LR-**), diagnostic odds ratio (**DOR**), and Youden’s Index (**YI**) which were based upon true positive (**TP**), false positive (**FP**), true negative (**TN**), and false negative (**FN**) diagnostic test accuracy metrics. Of the five reviewed articles which did not include true positive, false positive, true negative, and false negative diagnostic test accuracy metrics, the two independent reviewers retrieved TP, FP, TN, and FN data from primary studies and calculated the SE, SP, PPV, NPV, LR+, LR-, DOR, and YI.

##### Analysis of degree of overlap

A citation matrix was generated to calculate the CCA. This analysis classifies the degree of overlap as “slight” (zero to five percent), “moderate” (six to ten percent), “high” (11 to 15 percent) or “very high” overlap (greater than 15 percent).^15^ The formula used is CCA = (N-r) / (rc-r) where “N” is number of included primary studies, “r” equals the number of index publications, and “c” includes the number of reviews.

##### Estimation of common effect size

The primary aim of this umbrella review was to facilitate a straightforward comparison of the effects across various factors examined by utilizing a consistent measure of effect size. Since systematic reviews and meta-analyses incorporate different measures based on the design and analytical methods of the included studies, it was important to establish a common effect size to enable an overall comparison. To summarize from the included studies the effect size, mean, standard deviation, and confidence intervals were calculated for prevalence in percent, SE in percent, SP in percent, PPV in percent, NPV in percent, LR+, LR-, DOR, and YI.

#### Quality Appraisal, Risk of Bias Assessment, and Certainty of Evidence Assessment

The quality appraisal was conducted with the AMSTAR 2 tool, which enables the assessment of systematic reviews of randomized and non-randomized studies of healthcare interventions. The tool contains 16 domain items to assess the quality of included systematic reviews.^16^ The domain questions are designed so that a “yes” answer denotes a positive result and when information needed was present in the study. The “no” answer denotes when no information is available to rate. The “partial yes” answer denotes when partial information is provided in the article. The tool provides an overall rating based on weaknesses in critical domains.^16^

The risk of bias (**RoB**) assessment is included in two of the domain items of the AMSTAR 2 tool.^16^ The RoB from individual primary studies and from the interpretation of the results of the review are, respectively, included in domain “item 9” and “item 13” shown in Table 2. The risk of publication bias assessment utilized funnel-plot analytic methodology which is based on evidence for small-study effect.

The certainty of evidence was based upon the ten criteria Fusar-Poli guidance for umbrella reviews, which include 1) ensure that the umbrella review is really needed, 2) prespecify the protocol, 3) clearly define the variables of interest, 4) estimate a common effect size, 5) report the heterogeneity and potential biases, 6) perform a stratification of the evidence, 7) conduct sensitivity analyses, 8) report transparent results, 9) use appropriate software, and 10) acknowledge the limitations.^17^

## RESULTS

### Data Analysis and Results of Synthesis

#### Literature Search results and Study Selection

The initial literature search yielded 205 results, with 203 identified through database-search and two identified by hand-search. After removal of duplicates, 129 reviews were screened based on title and abstract, which resulted in exclusion of 98 studies. The remaining 31 reviews were read in full and 26 were excluded, which resulted in a total of five reviews^18–22^ included in this umbrella review and referred to as De Luca Canto *et al*, Michelet *et al*, Incerti Parenti *et al*, Patel *et al*, and Wu *et al* (Figure 1).

**Figure 1.**
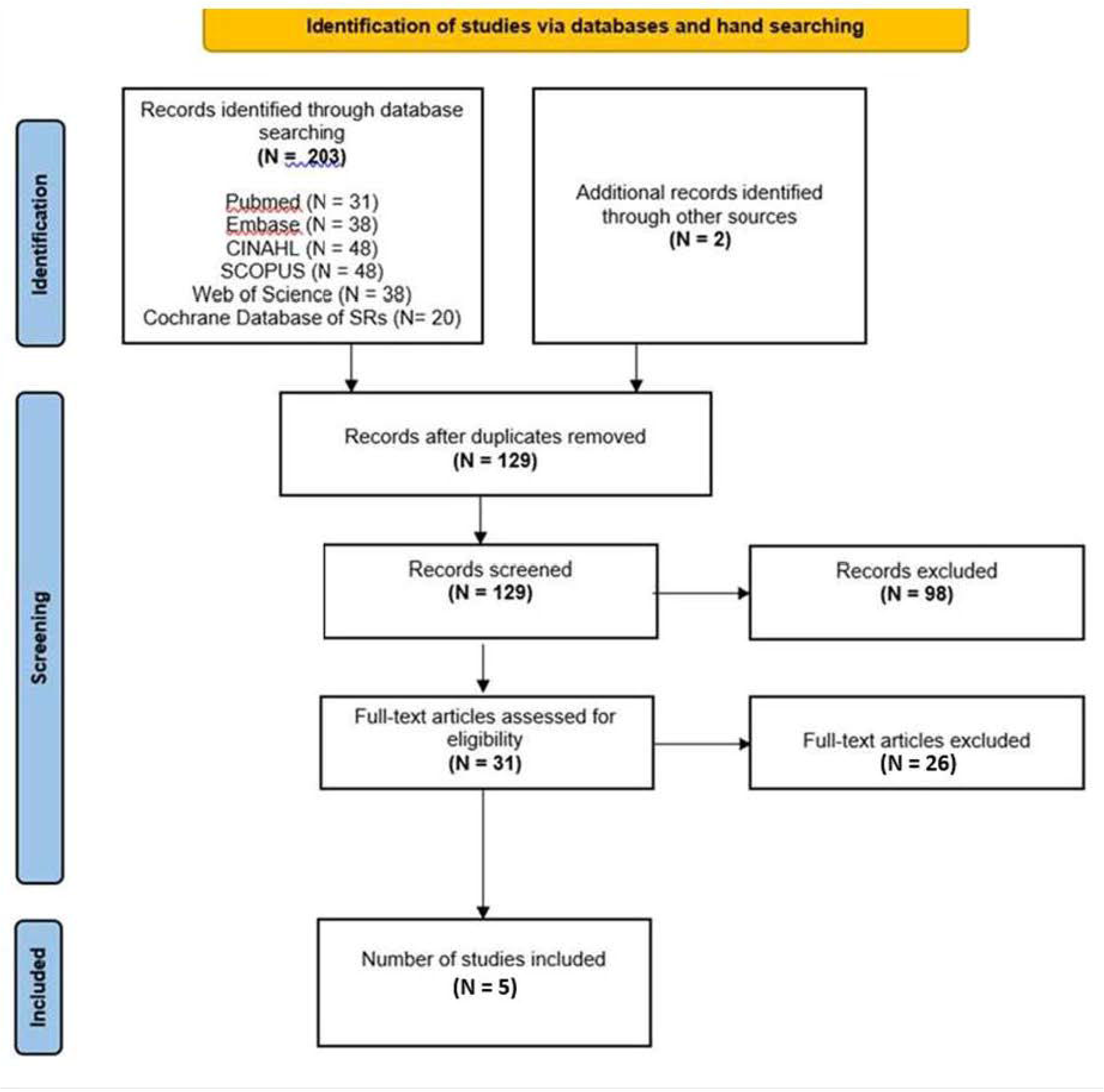
PRISMA flow diagram of identification and selection of included reviews.

#### Study characteristics

As shown in Table 1, one review was published in 2014 (Canada), one in 2019 (France), two in 2020 (Taiwan and United Kingdom), and one in 2021 (Italy).^18–22^ The number of searched databases reported by the included reviews ranged from four to seven, with the search period end-year ranging from 2013 to 2020.^18–22^ No language restrictions were reported for searches in any of the five included reviews. The primary designs and number of studies in the reviews varied with three using diagnostic testing accuracy with 11, 13, and 27 studies respectively^18–19,21^ one with observational designs with 13 quantitative studies^20^, and one using non-randomized clinical trials with 39 observational studies.^22^ The primary research question for all five reviews were based upon the accuracy of questionnaires for the PSG outcomes for the diagnosis of SRBDs in children, included the PSQ and PSQ-derivatives as the indicator, included PSG outcomes of AHI as the comparator, included the Quality of Diagnostic Accuracy Studies (**QUADAS**)^23^ as the assessment tool, and utilized meta-analysis methodology.^18–22^ Four of the five reviews shared similar conclusions with De Luca Canto *et al*. suggesting that dentists should use the PSQ as a screening tool to identify pediatric SRBDs; Michelet *et al* indicating that the PSQ-derived “SRBD scale has acceptable accuracy in detecting patients with” obstructive sleep apnea syndrome (**OSAS**); Incerti Parenti *et al* stating that the PSQ-derived SRBD “performed best and showed the highest sensitivity using the currently accepted diagnostic threshold for pediatric sleep apnea (AHI>=1); and Wu *et al* stating that the PSQ yielded the highest screening mild pediatric OSAS. ^18–20,22^ Only Patel *et al* indicated that the PSQ-derived SRBD scale and three other questionnaires could not be “considered as surrogates for PSG when diagnosing OSAS.^21^

**Table 1.**
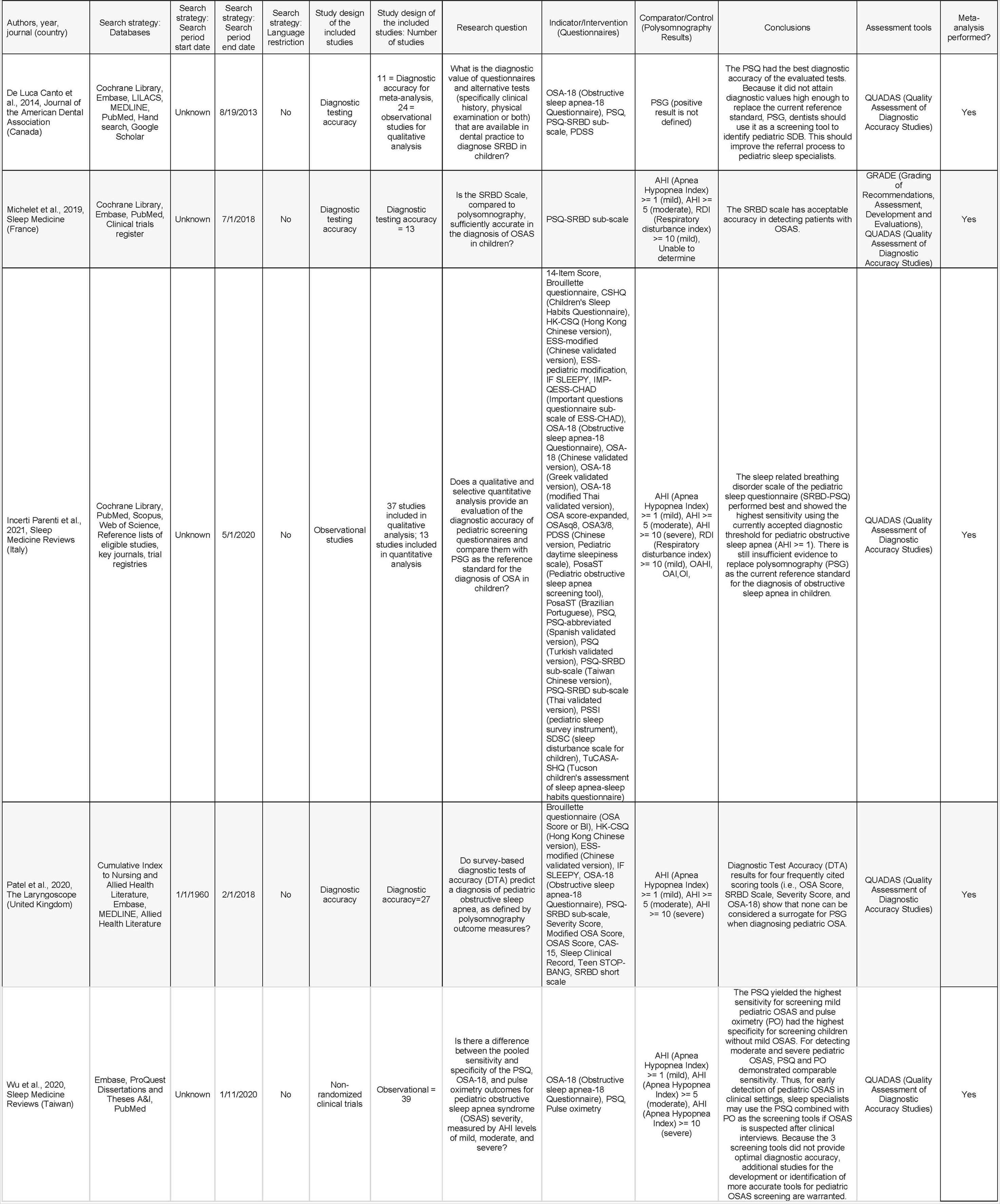
Characteristics of Included Articles.

**Table 2.**
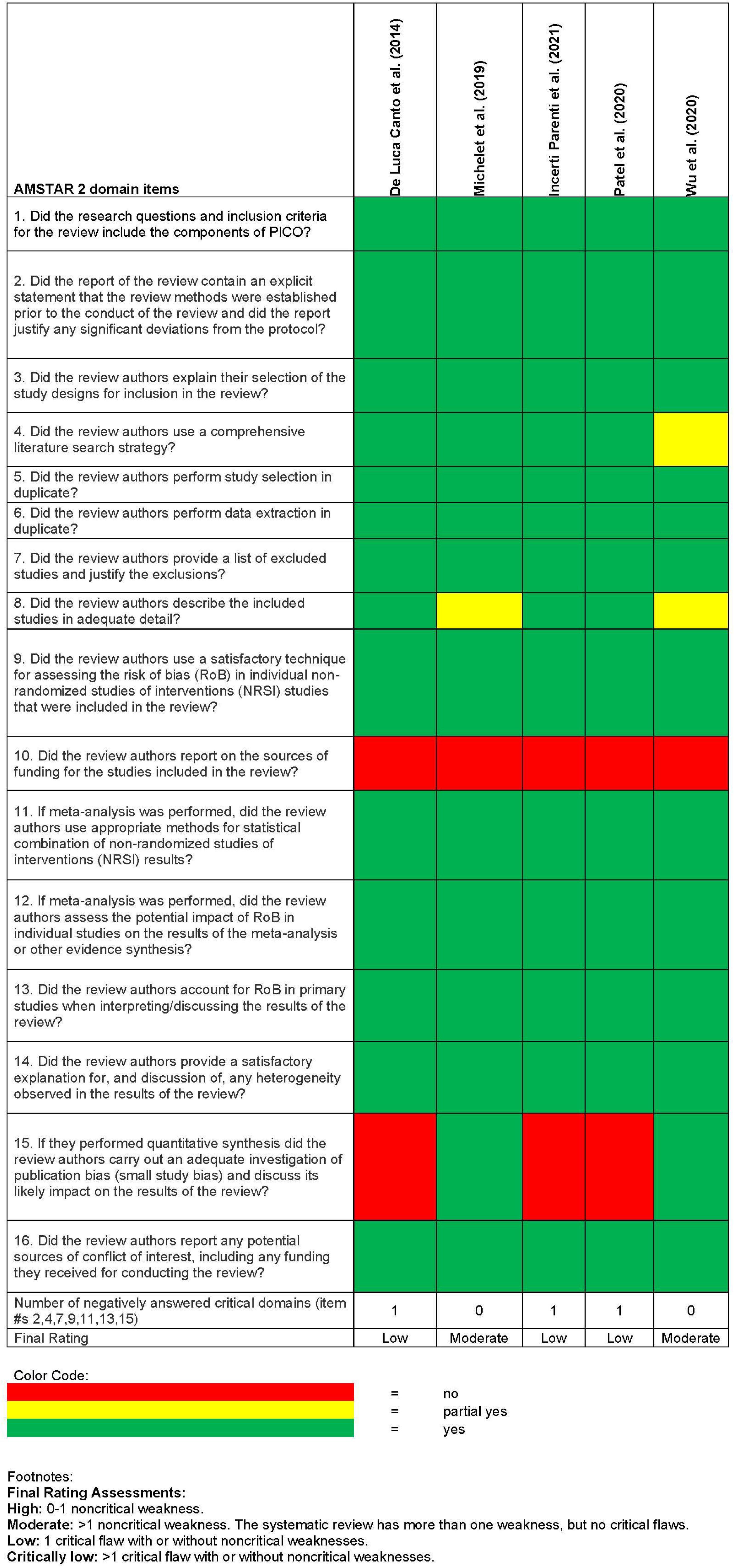
AMSTAR assessment.

#### Overlap of studies

Figure 2 shows the results of the CCA analysis. With five studies meeting the inclusion criteria, including a total of 21 index studies which focused on the PSQ as a screening tool for sleep disordered breathing, the degree of overlap between the reviewed studies was 26 percent.

**Figure 2.**
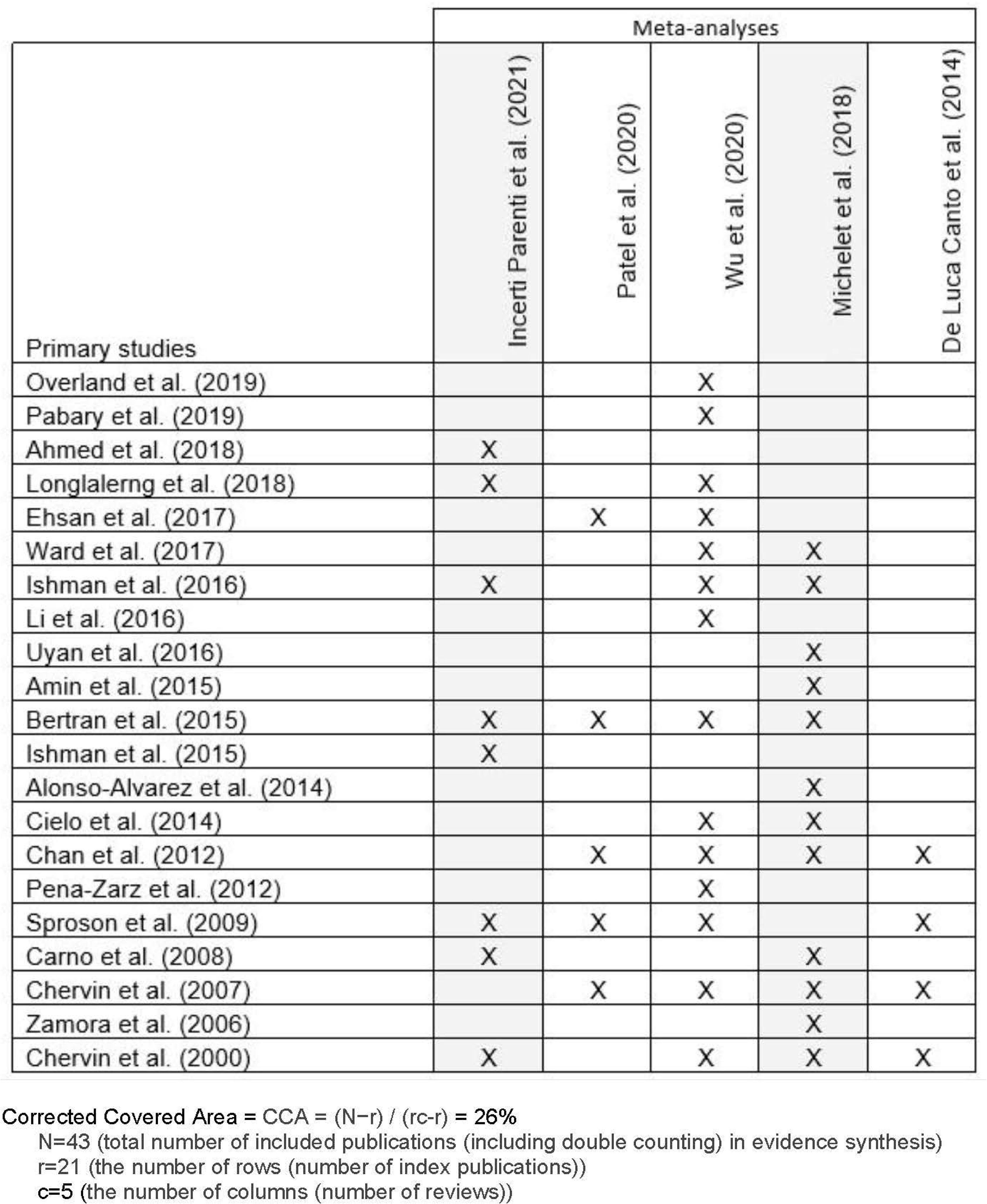
Corrected Covered Area (CCA)

#### Meta-analysis summary

A tabular summary of the meta-analysis of the diagnostic test accuracy metrics shows the point-estimate mean, standard deviation (**SD**), and Confidence Interval (**CI**) all diagnostic testing accuracy metrics (Table 3). The prevalence among children was 50.58% percent (SD=23.28, 95% CI [44.29, 56.87]). The average SE and SP of the diagnostic tool were, respectively, 74.97 (SD=14.78, 95% CI [70.97, 78.96]) and 49.68 percent (SD=22.42, 95% CI [43.62, 55.74]). The average PPV was 58.16 percent (SD=26.52, 95% CI [51.23, 65.08]). The average NPV was 62.24 percent (SD=21.65, 95% CI [56.39, 68.09]). The average LR+ was 2.08 (SD=1.70, 95% CI [1.62, 2.54]) and the average LR- was 0.57 (SD=0.33, 95% CI [0.48, 0.66]). The average DOR was 6.94 (SD=9.66, 95% CI [4.30, 9.58]) and average YI was 0.25 (SD=0.23, 95% CI [0.24, 0.25]). The average number of true positive was 35.22, false positive was 29.20, false negative was 12.13, and true negative was 24.78.

**Table 3.** Diagnostic Testing Accuracy Metrics, with confidence intervals.

| Overall |  |  |  | De Luca<br>Canto et<br>al. (2014) | Michelet<br>et al.<br>(2018) | Incerti<br>Parenti et<br>al. (2021) | Patel et al.<br>(2020) | Wu et al.<br>(2020) |  |
| --- | --- | --- | --- | --- | --- | --- | --- | --- | --- |
| N<br>(primary studies) | 55 |  |  | 2 | 13 | 12 | 5 | 23 |  |
| n (total subjects) | 5697 |  |  | 183 | 1162 | 861 | 508 | 2983 |  |
| Variable<br>(mean (SD)) | Mean<br>(SD) | 95% CI<br>Lower<br>Bound | 95% CI<br>Upper<br>Bound |  |  |  |  |  | p-value |
| Prevalence (%) | 50.58<br>(23.28) | 44.29 | 56.87 | 56.00<br>(41.01) | 51.91<br>(25.32) | 58.30<br>(21.24) | 47.00<br>(19.46) | 46.11<br>(23.44) | 0.671 |
| Sensitivity (%) | 74.97<br>(14.78) | 70.97 | 78.96 | 65.50<br>(21.92) | 68.55<br>(19.28) | 78.56<br>(10.45) | 73.18<br>(13.88) | 77.93<br>(13.14) | 0.295 |
| Specificity (%) | 49.68<br>(22.42) | 43.62 | 55.74 | 71.20<br>(22.35) | 57.17<br>(21.35) | 51.29<br>(24.30) | 41.84<br>(25.74) | 44.44<br>(20.66) | 0.268 |
| Positive predictive<br>value (%) | 58.16<br>(25.62) | 51.23 | 65.08 | 63.50<br>(47.66) | 60.09<br>(26.57) | 65.96<br>(23.09) | 54.28<br>(18.77) | 53.37<br>(26.97) | 0.715 |
| Negative<br>predictive<br>value (%) | 62.24<br>(21.65) | 56.39 | 68.09 | 59.65<br>(21.14) | 61.59<br>(20.01) | 58.76<br>(18.64) | 59.20<br>(25.45) | 65.32<br>(24.51) | 0.929 |
| Positive likelihood<br>ratio | 2.08<br>(1.70) | 1.62 | 2.54 | 3.65<br>(3.61) | 2.37<br>(1.91) | 2.31<br>(1.95) | 1.46<br>(0.78) | 1.79<br>(1.42) | 0.468 |
| Negative<br>likelihood ratio | 0.57<br>(0.33) | 0.48 | 0.66 | 0.55<br>(0.50) | 0.59<br>(0.33) | 0.51<br>(0.26) | 0.80<br>(0.42) | 0.55<br>(0.34) | 0.582 |
| Diagnostic Odds<br>Ratio | 6.94<br>(9.66) | 4.30 | 9.58 | 14.85<br>(19.30) | 8.55<br>(11.65) | 8.78<br>(12.00) | 2.96<br>(3.51) | 5.17<br>(6.76) | 0.457 |
| Youden's Index | 0.25<br>(0.23) | 0.24 | 0.25 | 0.37<br>(0.44) | 0.26<br>(0.26) | 0.30<br>(0.25) | 0.15<br>(0.22) | 0.22<br>(0.20) | 0.704 |
| True positive (n) | 35.22<br>(32.39) | 26.38 | 44.06 | 35.0<br>(29.69) | 32.46<br>(20.10) | 31.50<br>(16.09) | 34.60<br>(18.37) | 39.05<br>(46.33) | 0.970 |
| False positive (n) | 29.20<br>(37.31) | 19.11 | 39.29 | 17.50<br>(21.92) | 19.77<br>(20.98) | 15.33<br>(11.66) | 31.20<br>(24.28) | 42.35<br>(51.27) | 0.237 |
| False negative (n) | 12.13<br>(15.80) | 7.86 | 16.40 | 13.50<br>(0.71) | 11.77<br>(10.80) | 8.50<br>(3.99) | 10.80<br>(3.12) | 14.39<br>(22.94) | 0.893 |
| True negative (n) | 24.78<br>(26.66) | 17.58 | 31.99 | 26.00<br>(21.21) | 25.31<br>(27.22) | 12.83<br>(8.98) | 25.00<br>(18.86) | 30.57<br>(33.19) | 0.049 |

Selective forest-plots are visualized, respectively, to illustrate magnitude and level of heterogeneity in Figure 3(a), 3(b), 3(c), 3(d), and 3(e) for the diagnostic testing accuracy metrics for SE, SP, PPV, NPV, and DOR. These five forest-plots show the point-estimates for each metric and low levels of heterogeneity amongst the five reviewed studies. The point estimate for SE is relatively high at 74.97 percent, SP is moderate at 49.68 percent, PPV is moderate at 58.16 percent, and NPV is moderate at 62.24 percent. The point-estimate for the DOR is positive at 6.94.

**Figure 3(a), (b), (c), (d), (e).**
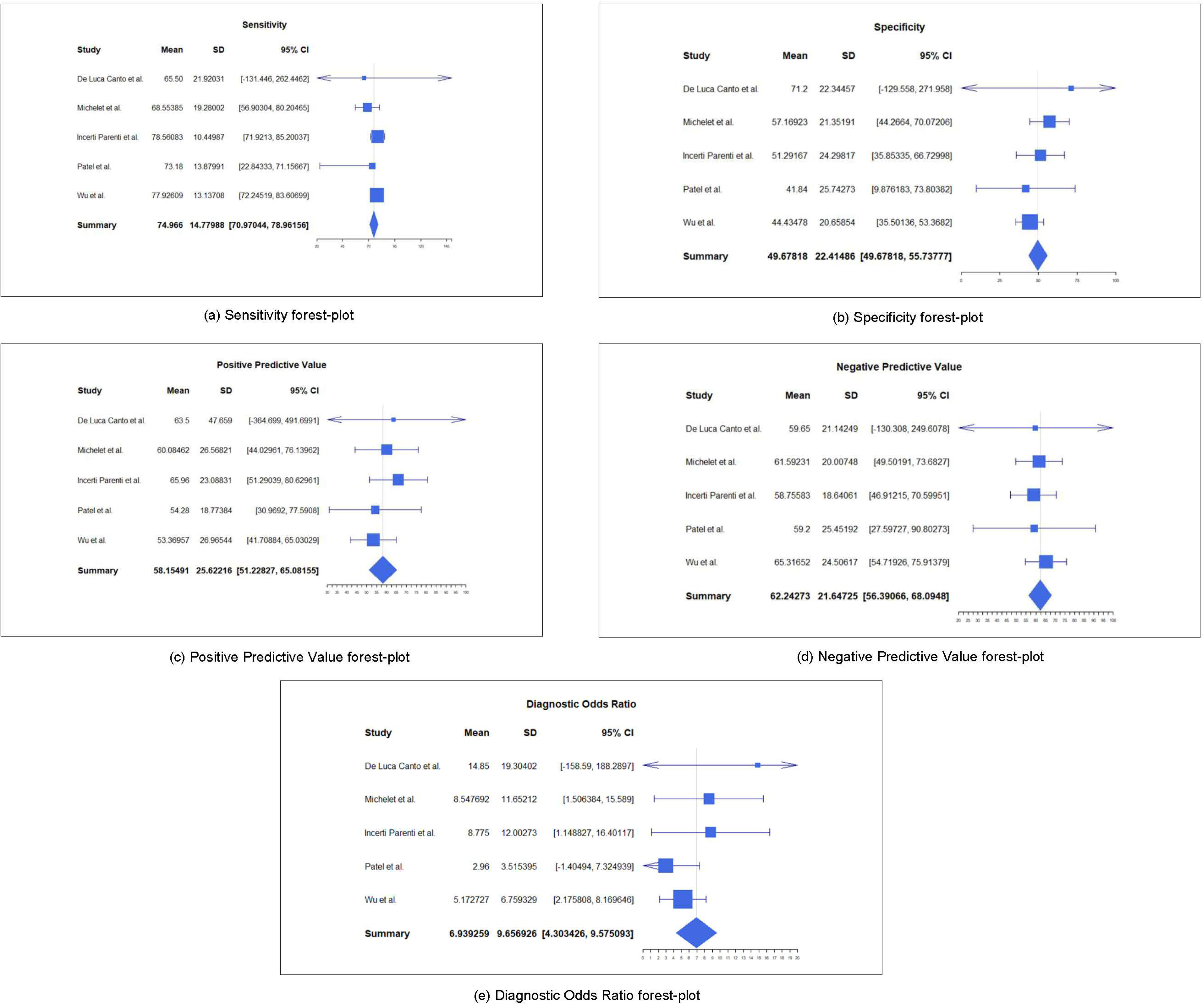
Forest Plots by Diagnostic Testing Accuracy metric.

#### Methodological Quality - Quality appraisal, RoB, Heterogeneity of the Data, and COE

The results of the AMSTAR 2 quality appraisal of the 5 reviews are presented in Table 2. Three studies (60 percent) were classified as being of low quality per the AMSTAR 2 rating scale, and two studies (40 percent) were classified as being of moderate quality.

Three of the reviews (60 percent)^18,20,21^ did not conduct the critical investigation of publication bias (item 15), which lowered the quality assessment. Additionally, the item that was most frequently omitted in the reviews was “item 10,” the non-critical sources of funding for five (100 percent) of the studies.^18–22^

#### Risk of Bias

Table 2 shows five (100 percent) of the included reviews had “yes” responses for “item 9” and “item 13” in the AMSTAR 2 assessment. Item 9 asks, *“*Did the review authors use a satisfactory technique for assessing the risk of bias (RoB) in individual studies that were included in the review?” *and item 13, “*Did the review authors account for RoB in primary studies when interpreting/discussing the results of the review?^18,19,20,21,22^” ***Publication Bias***

Table 2 shows two (40 percent) of the included reviews had “yes” responses for “item 15” in the AMSTAR 2 assessment which yielded “Moderate” ratings and asked, *“*If they performed quantitative synthesis did the review authors carry out an adequate investigation of publication bias (small study bias) and discuss its likely impact on the results of the review?^19,22^” The three other studies received “no” responses which constituted critical flaws and downgraded their ratings to “Low.^18,20,21^”

Additionally, the primary studies for all five reviewed studies were quantitatively compiled and analyzed.

Figure 4 shows the funnel-plot which demonstrates a low risk of publication bias since the standard error and standardized mean difference for all five reviewed studies fall under the funnel boundaries.

**Figure 4:**
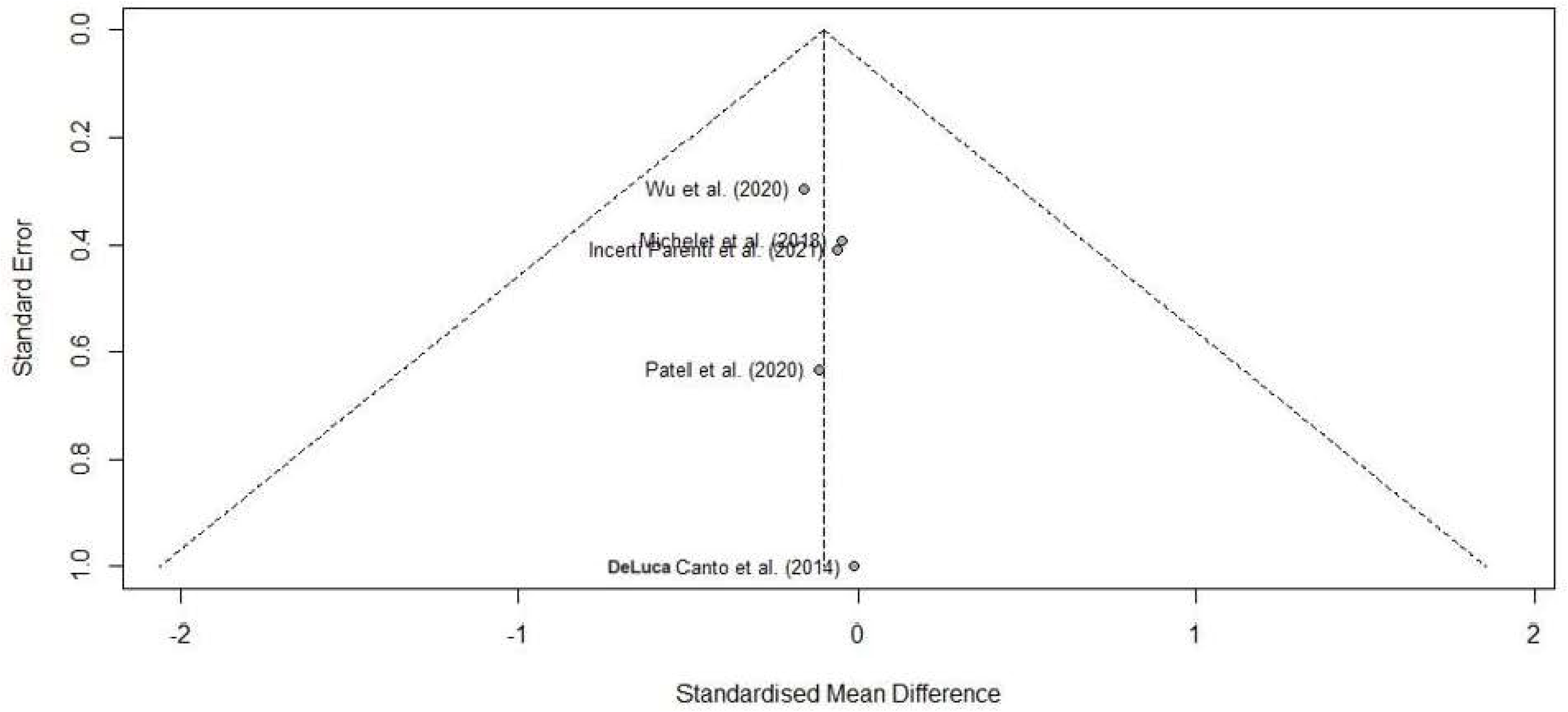
Publication bias funnel-plot for clustered five included reviews.

#### Heterogeneity of Data

Table 2 also shows five (100 percent) of the included reviews had “yes” responses for “item 14” in the AMSTAR 2 assessment, which asked, *“*Did the review authors provide a satisfactory explanation for, and discussion of, any heterogeneity observed in the results of the review?^18–22^”

The primary studies for all five reviewed studies were quantitatively compiled and analyzed. Figure 5 shows very little heterogeneity of the standardized mean difference, with an I^2^-statistic equal to zero percent.

**Figure 5.**
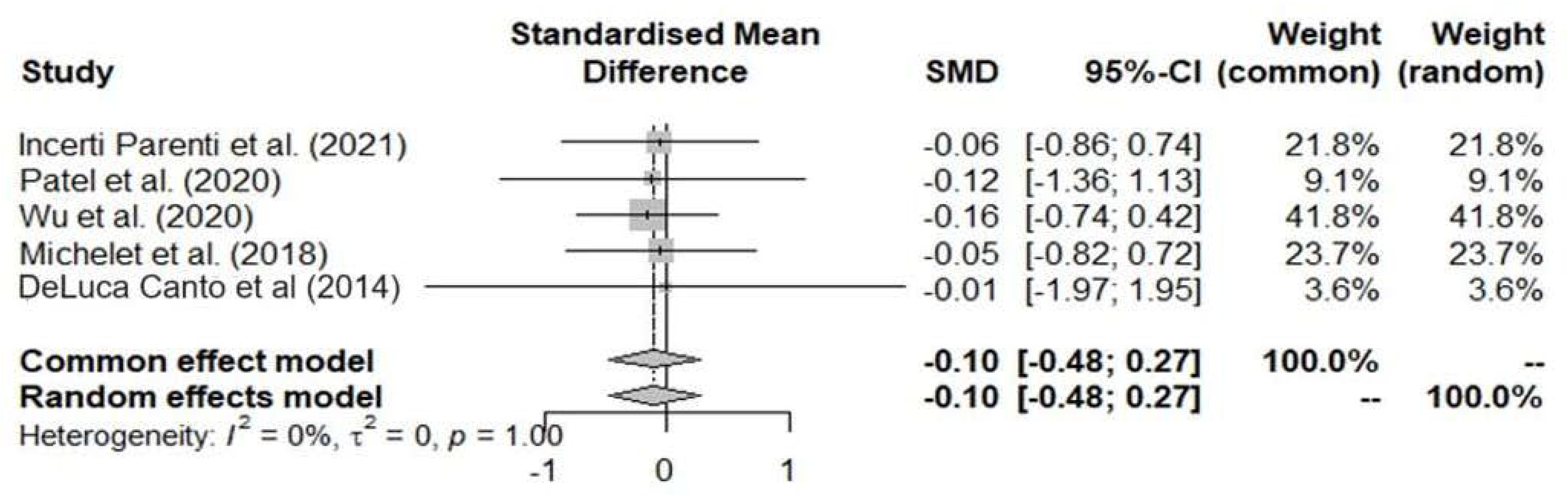
Heterogeneity analysis for clustered five included reviews.

#### Certainty of Evidence

The certainty of evidence summary was based upon and fulfills the ten criteria from the Fusar-Poli model listed below.^17^

1. *Ensure that the umbrella review is really needed:*

*a.* Neither the March 2022 nor June 2023 literature search yielded previously reported umbrella reviews with the same research purpose and PICO question as this present study (Figure 1).
2. *Prespecify the protocol*:

*a.* The present study was registered with the Prospero register for systematic reviews and was assigned the identification number CRD42023393232.
3. *Clearly define the variables of interest:*

*a.* The variables of interest were defined as the diagnostic test accuracy metrics for prevalence, sensitivity, specificity, positive predictive value, negative predictive, positive likelihood ratio, negative likelihood ratio, diagnostic odds ratio, and Youden’s Index (Table 3).
4. *Estimate a common effect size:*

*a.* The common effect size was calculated for the selected diagnostic test accuracy metrics for mean point estimate, standard deviation, and 95 percent confidence intervals with lower and upper bounds (Table 3).
5. *Report the heterogeneity and potential biases:*

*a.* Heterogeneity was calculated utilized the I^2^ statistic, which equaled zero percent with an interpretation of low heterogeneity (Figure 5).
6. *Perform a stratification of the evidence:*

*a.* Stratification of the evidence utilized the classification method defined as, “Class I (convincing) when the number of cases > 1,000; I^2^ less than 50 percent; 95 percent interval excluding the null, no small-study effects (publication bias) and no excess significance bias.” The total number of cases from the primary studies equaled 5,967 subjects and fulfilled the I^2^ statistic, 95 percent confidence interval excluding the null.
7. *Conduct sensitivity analyses:*

*a.* Of the primary studies 69% (113/164) and 31% (51/164) were respectively designed as observational and diagnostic testing accuracy studies, for which association does not necessarily imply causation and limits the ability to analyze sensitivity.
8. *Report transparent results:*

*a.* With the aim of transparency, this study intended to summary the evidence clearly by way of table and figures. Specifically, the study included data from the primary studies to generate the comparative diagnostic testing accuracy metrics in tabular format and subsequent forest-plots and funnel-plot analyses.
9. *Use appropriate software:*

*a.* All analyses were performed in R version 4.0.3 (The R Foundation for Statistical Computing, Vienna, Austria).^24^
10. *Acknowledge the limitations:*

*a.* Our study has several limitations with the AMSTAR 2 assessments demonstrating low to moderate quality related to publication bias and the CCA calculated at 26%, as detailed in the discussion section.

## DISCUSSION

### Interpretations of Results

#### Key Findings and Relationship to the Hypothesis

The key findings from this umbrella review found the PSQ to be an accurate and reliable tool as a tool for dentists to routinely screen for SRBDs in pediatric patients. Although the AMSTAR 2 quality of evidence ranges from low to moderate, the PSQ was shown to be useful as a screening tool for pediatric SRBDs as modeled by Trevethan.^12^ The model shows with moderate levels of sensitivity (74.97%), specificity (49.68%), positive predictive value (58.18%), and negative predictive value (62.24%), PSQ screening is useful since SRBDs may not progress quickly or benefit from early referral and no harm is likely to be done to children by screening them for SRBDs even if this condition is not present. Hence, there is sufficient evidence to support the hypothesis that the PSQ is sufficiently accurate as a screening tool for the diagnosis of SRBDs in children.

### Quality of the included systematic reviews and meta-analyses

The AMSTAR 2 tool was used for the quality appraisal of the included studies. According to this tool, a “high” quality study provides an accurate summary of results while a “low” quality study may not be as accurate. In this umbrella review, three of the studies were of low quality while two studies were of moderate quality. The primary factor for the low quality studies was that publication bias was not investigated.

However, the quantitative analysis showed that there was low risk of publication bias from the funnel-plot analysis and low heterogeneity, with sufficiently high diagnostic testing accuracy metrics, for SE, SP, PPV, NPV, and DOR from the meta-analysis data.

### Strengths

This review has systematically summarized the PSQ as a tool to screen for sleep apnea in the pediatric population from existing systematic reviews. Several criteria can be considered a strength. One strength was a comprehensive search strategy was performed in multiple electronic databases to avoid missing relevant systematic reviews; second, two reviewers independently assessed the quality of evidence. AMSTAR 2 was used as a critical assessment tool for systematic reviews in this umbrella review. Another strength was the extraction of data from the 55 primary articles with nearly 6,000 subjects. Also, based upon the Fusar-Poli model, this study fulfilled the ten criteria for certainty of evidence.

### Limitations

The study had a few limitations, which included the AMSTAR 2 assessments with low to moderate quality for the 5 included reviews and three of the studies not investigating publication bias, which is a critical flaw. Another limitation is the high CCA value with 26 percent of index studies appeared multiple times across the five included reviews, which increased the weighting of the results.

### Principal findings and Implications for Practice, Policy, and Future Research

Based upon the Trevethan^12^ model there is sufficient evidence to support the PSQ as a useful screening tool for practice, policy, and future research.

The AAPD guidance, which states that the PSQ is “not sensitive enough to detect presence or severity of OSA,” is based upon one Norwegian study with 100 children ages two to six years old who were referred for adenotonsillectomy surgery and found the respective diagnostic test accuracy metrics prevalence for children with AHI ≥ 1 as follows: prevalence (87 percent), SE (75.6 percent), SP (45.5 percent), PPV (91.6 percent), and NPV (19.2 percent).^25^ However, the interpretation of this study did not take into account the interaction between SE, SP, PPV, and NPV, as described by Trevethan, who states that, “a certain percentage of false positive outcomes (i.e., a moderate SE level of 75.6 percent) might not be objectionable if follow-up tests are inexpensive, easily and quickly performed, and not stressful for clients (e.g., referral to a physician for suspected pediatric sleep-related breathing disorders). In addition, false positive screening outcomes might be quite permissible if no harm is likely to be done to clients in protecting them against a target condition even if that condition is not present.^12^” As an example, children who are mistakenly told that they may have a sleep-related breathing disorder, are likely to receive benefits from a referral to their primary care provider (**PCP**) and encouraged to practice preventive recommendations for healthy diet, regular exercise, and air quality hygiene.

The interaction and interpretation between prevalence, SE (74.97 percent), SP (49.68 percent), PPV (58.16 percent), and NPV (62.24 percent) also applies to the results for this present study. As described by Power, the “Sensitive test when Negative rules OUT the disease” (**SNOUT**) rule of thumb can be used for a screening test with high SE, low SP, moderate PPV and NPV levels, which exceed the threshold for action (i.e., referral of children with suspected sleep-related breathing disorder from a positive PSQ screening to their PCP for assessment).^11^

With the high prevalence of SRBDs in children in this study (50.58%) and limited nation-wide capacity for children to undergo polysomnography testing, it is clear that the PSQ should be utilized by dentists to adhere to the AAP and AAPD guidelines for screening and referring children with suspected SRBDs.

Future studies should include more randomized controlled trials to identify other tests which better refine the prediction of pediatric SRBDs. Examples of such tests include home-sleep tests, app-based tests for cell-phones, wrist-watches, finger-rings, and artificial intelligence-based tests for children based upon biometric data for sleep quality.

## CONCLUSIONS

Based upon the results of this study, the following conclusions can be made:

1. Dentists should adhere to the AAPD guidelines and know that there is low to moderate evidence to support the utilization of the PSQ as a sufficiently accurate screening tool for the diagnosis of SRBDs in children, followed by an appropriate referral to a physician for comprehensive assessment and management.

## Data Availability

All data produced in the present study are available upon reasonable request to the authors.

## AKNOWLEDGMENTS

The authors extend appreciation and acknowledgments for funding to the Hansjorg Wyss Department of Plastic Surgery, NYU Langone Hospitals, New York, NY and to Sallie Yassin, MS who provided valuable support for the data analysis.

